# Specificity of cortical area and thickness as biomarkers for comorbid internalizing and externalizing mental disorders in pre-adolescence

**DOI:** 10.1101/2022.04.24.22273790

**Authors:** Nanyu Kuang, Zhaowen Liu, Gechang Yu, Kai Zhang, Xinran Wu, Ben Becker, Huaxin Fan, Jiajia Zhao, Jujiao Kang, Guiying Dong, Xingming Zhao, Jianfeng Feng, Barbara J. Sahakian, Trevor W. Robbins, Gunter Schumann, Lena Palaniyappan, Jie Zhang

## Abstract

**Background:** Comorbidity is the rule rather than the exception for childhood and adolescent onset mental disorders, but we cannot predict its occurrence and do not know the neural mechanisms underlying comorbidity. We investigate if the effects of comorbid internalizing and externalizing disorders on anatomical changes represent a simple aggregate of the effects on each disorder, and if comorbidity-related cortical surface changes relate to a distinct genetic underpinning.

**Methods:** We studied the cortical surface area (SA) and thickness (CT) of 11,878 preadolescents (9-10 years) from the Adolescent Brain and Cognitive Development Study. Linear mixed models were implemented in comparative and association analyses among internalizing (Dysthymia, Major Depressive Disorder, Disruptive Mood Dysregulation Disorder, Agoraphobia, Panic Disorder, Specific Phobia, Separation Anxiety Disorder, Social Anxiety Disorder, Generalized Anxiety Disorder, Post-Traumatic Stress Disorder), externalizing diagnostic groups (Attention-Deficit/Hyperactivity Disorder, Oppositional Defiant Disorder, Conduct disorder) a group with comorbidity of the two and a healthy control group. Genome-wide association analysis and cell type specificity analysis were performed on 4,716 unrelated European participants from this cohort.

**Results:** Reduced cortical surface area but increased thickness occurs across patient groups when compared to controls. Children with comorbid internalizing and externalizing disorders had more pronounced areal reduction than those without comorbidity, indicating an additive burden. In contrast, cortical thickness had a non-linear effect with comorbidity: the comorbid group had no significant CT changes, while those patient groups without comorbidity had significant thickness increases. Distinct biological pathways were implicated for regional SA and CT changes. Specifically, CT changes were associated with immune-related processes implicating microglia, while SA-related changes related mainly to excitatory neurons.

**Conclusions:** The emergence of comorbidity across distinct clusters of psychopathology is unlikely to be a simple additive neurobiological effect. Distinct risk-adaptation processes, with unique genetic and cell-specific factors may underlie SA and CT changes. Children with highest risk but lowest resilience, both captured in their developmental morphometry, develop a comorbid illness pattern.

## Introduction

Adolescence is a vulnerable period for gray matter maturation and many psychiatric disorders of adulthood begin at the preadolescent stage (1–3). The preadolescent disorders can be broadly classified into internalizing and externalizing disorders, with a high degree of comorbidity between them (4). For example, anxiety disorders (internalizing) are often comorbid with externalizing disorders such as attention deficit hyperactivity disorder (ADHD) (5–8) or conduct disorder (CD) (9); Oppositional defiant disorder (ODD, externalizing) being comorbid with anxiety or depression (internalizing) (10); This pattern is especially common in pre-adolescent period (11), during which the prevalence of comorbidity is greater than that of individual groups of disorders (12). This pattern of comorbidity indicates diminished response to conventional treatments as well as poor functional outcomes (13). Furthermore, the pattern of comorbidity often emerges over time, and not identifiable at the outset, at the time of first presentation, precluding early interventions aimed at comorbidity. Despite this significant burden resulting from comorbidity, it is not clear if ywe can identify unique markers for comorbidity at the outset. We also do not know if comorbidity results from additive effect of disorder-specific mechanisms (shared) or arise from processes that are unique to the comorbid trajectory.

Transdiagnostic neruoimaging biomarkers have been identified, with the potential to track the vulnerability for psychiatric disorders even before overt clinical presentations occur (14, 15). Two MRI-based markers of cortical morphology with distinct genetic basis and developmental trajectory (16, 17) are surface area (SA) and cortical thickness (CT). According to the radial unit hypothesis, expansion of SA is driven by the proliferation of neural progenitor cells and tangential migration, while CT is related to the number of neurogenic divisions and radial migration (18). Several studies indicate that internalizing and externalizing disorders have unique neurodevelopmental patterns reflected by their CT and SA alterations (19). Some studies report opposing changes in CT in internalizing and externalizing disorders (e.g., anxiety relates to higher CT in the prefrontal cortex (PFC) and precentral gyrus (20) while ADHD relates to reduced CT in the PFC and precentral regions (21).

A previous examination of the ABCD cohort found no association between general psychopathology (internalizing and externalizing symptoms) with CT. However, comorbidity was not specifically studied in this analysis (22). In contrast, SA was found to be correlated with general psychopathology. This finding was validated in another independent cohort (mean age 10.6 years old) (19). Comorbidity of internalizing and externalizing disorders may have different underlying effects: On the one hand, CT and SA changes may be the result of additive influences of both disorders, with comorbid children exhibiting both patterns when compared to the healthy group. On the other hand, if a distinct rather than additive impairment results in comorbidity, we are more likely to see unique patterns of CT in comorbid cases (specific effect). Further, CT and SA are under the influence of distinct sets of genes and biological processes (23–25). Determining the unique contributions of CT and SA to comorbid internalizing and externalizing disorders could help uncover the developmental neurobiology of comorbidity. Ultimately, this may provide a reliable means for characterizing children who are likely to develop comorbidity for these two families of disorders.

In this study, we empirically test for the presence of additive (i.e. a simple aggregate effect) vs. unique morphometric patterns in children with internalizing and externalizing comorbidity using a large developmental cohort of preadolescent participants of the ABCD study (26). While we remain agnostic as to the presence of additive vs. specific effects for comorbidity, we anticipate a divergence between CT and SA, given their discordant genetic and maturational trajectories (27–30). Within this cohort, we selected the homogeneous group of unrelated European youths to perform a Genome-Wide Association Analysis (GWAS) and locate the genetic variants associated with regional SA and CT changes. This analysis was carried out in conjunction with a search for common genetic elements across the affected brain regions from ABCD study, and a determination of the brain cell type-specific expressions that shared maximum variance with the patterns of morphometric changes observed in the patient sample. Within a disorder family, high degree of similarities exist among individual disorders in terms of genetic heritability (31, 32) and neuroanatomical patterns (33). As a result, we only consider comorbidity between the larger diagnostic families, i.e., between internalizing and externalizing disorders (34). Children with though disorders were excluded due to the small sample size.

## Methods

### Definition of diagnostic families

Mental disorder diagnoses were determined using parent or guardian responses to the parent-reported computerized Schedule for Affective Disorders and Schizophrenia for School-Age Children for DSM-5 (K-SADS-5) (35). Lifetime (past or present) diagnoses of the 18 disorders were used (22, 36, 37). Based on the definition of broad diagnostic families adopted in recent studies (36, 38, 39), two broad diagnostic families including externalizing disorders (Attention-Deficit/Hyperactivity Disorder, Oppositional Defiant Disorder, Conduct disorder), internalizing disorders (Dysthymia, Major Depressive Disorder, Disruptive Mood Dysregulation Disorder, Agoraphobia, Panic Disorder, Specific Phobia, Separation Anxiety Disorder, Social Anxiety Disorder, Generalized Anxiety Disorder, Post-Traumatic Stress Disorder) were used in our analysis (Figure 1A). We excluded children with though disorders (Hallucinations, Delusions, Associated Psychotic Symptoms, Bipolar Disorder, Obsessive-Compulsive Disorder) due to the small sample size (n=347).

**Figure 1.**
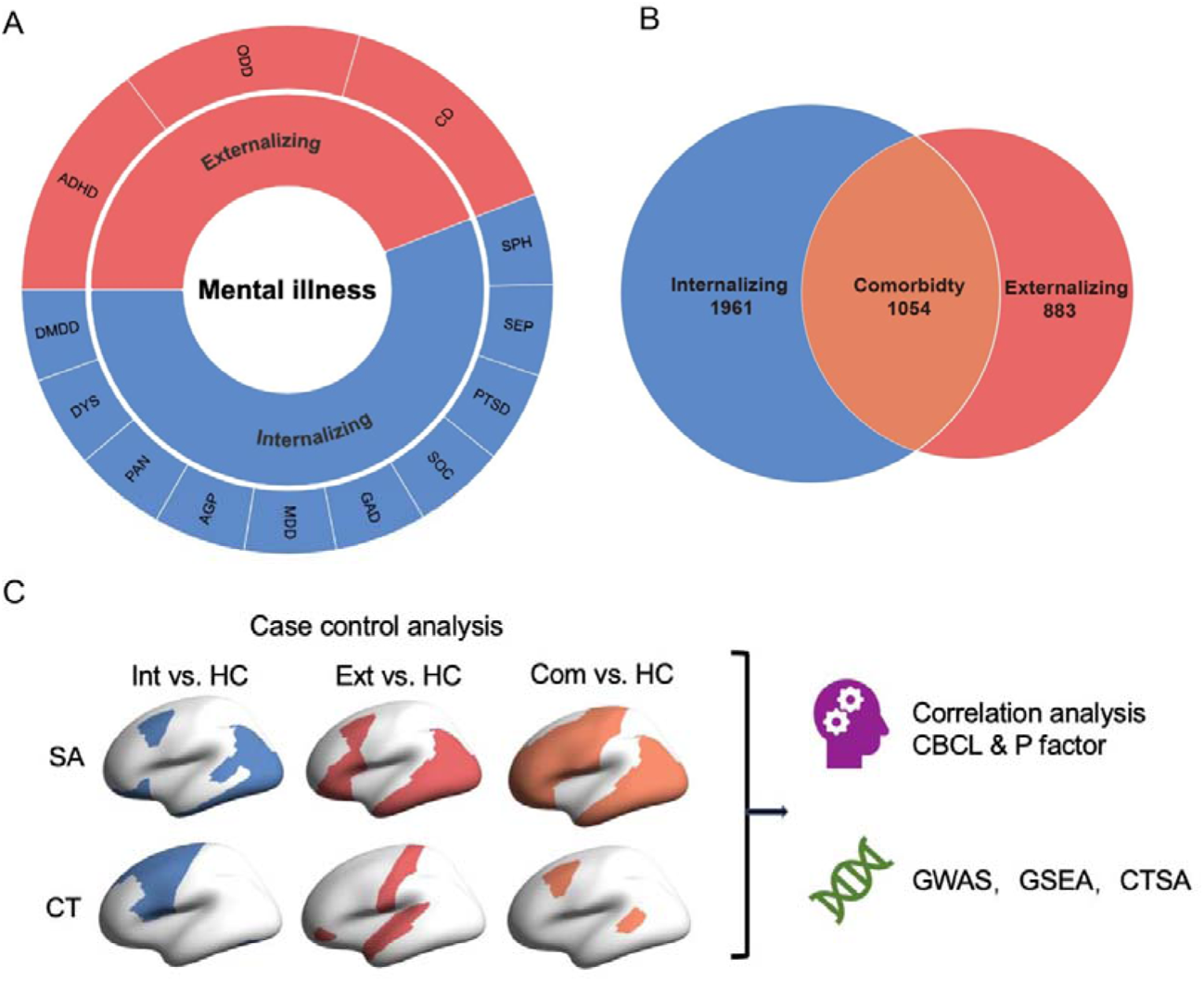
Components and comorbidity of externalizing and internalizing disorders. **(A)** 13 mental disorders (outer circle) were classified into two transdiagnostic categories (inner circle), i.e., externalizing and internalizing disorders. **(B)** Venn diagram depicting the overlap between the 2 transdiagnostic categories. Pure subsets of two transdiagnostic categories: externalizing disorder, red; internalizing disorder, blue. Comorbid between internalizing and externalizing disorders, orange. Children with thought disorder have been eliminated from membership of either external disorder or internal disorder. **(C)** An overview of all analysis. Abbreviations: ADHD, attention deficit hyperactivity disorder; CD, conduct disorder; ODD, oppositional defiant disorder; MDD, major depressive disorder; GAD, generalized anxiety disorder; SOC, social anxiety disorder; SEP, separation anxiety disorder; PTSD, post-traumatic stress disorder; AGP, agoraphobia; SPH, specific phobia; PAN, panic disorder; DYS, dysthymia; DMDD, disruptive mood dysregulation disorder; DEL, delusions; SA, surface area; CT, cortical thickness; Int, Internalizing disorders; Ext, Externalizing disorders; Com, Comorbid between internalizing and externalizing disorders; HC, healthy control; CBCL, Child Behavior Checklist; GWAS, Genome-wide association study; GSEA, Gene set enrichment analysis; CTSA, Cell type specificity analysis.

We consider 3 broader diagnostic groups: pure internalizing and externalizing disorders and their comorbidity, see Figure 1B. Healthy control preadolescents were those who did not have any mental disorders diagnoses (including unspecified or other specified disorders, eating disorders, Alcohol Use Disorder, Substance Related Disorder, Sleep Problems, Suicidal ideation or behavior and Homicidal ideation or behavior).

### Participants

Preadolescents aged 9-10 years (n=11,878) are recruited from 22 research sites across the USA from the Adolescent Brain Cognitive Development (ABCD) Study® (Release3.0, November 2020), which contains physical and mental health, cognition, genetic and neuroimaging data. Lifetime psychiatric diagnoses were determined using K-SADS-5. Demographic data of full ABCD sample was listed in Table 1. We consider four groups: externalizing, internalizing, comorbidities between internalizing and externalizing disorders and a healthy control group.

**Table 1.** Demographics of the three transdiagnostic categories (n=7570).

| Characteristic | Ext,<br>n = 883 | Int,<br>n = 1961 | Com,<br>n=1054 | HC,<br>n=3672 |
| --- | --- | --- | --- | --- |
| Age, mean (SD)<br>(months) | 119.36 (7.45) | 119.12 (7.58) | 119.19 (7.54) | 119.11 (7.48) |
| Gender, n (%) |  |  |  |  |
| Male | 598 (67.72%) | 899 (45.84%) | 634 (60.15%) | 1729 (60.79%) |
| Race, n (%) |  |  |  |  |
| White | 515 (58.32%) | 1131 (57.67%) | 613 (58.16%) | 1911 (52.04%) |
| Black | 122 (13.82%) | 232 (11.83%) | 129 (12.24%) | 506 (13.78%) |
| Hispanic | 153 (17.33%) | 386 (19.68%) | 166 (15.75%) | 794 (21.62%) |
| Asian | 7 (0.79%) | 27 (1.38%) | 10 (0.95%) | 97 (2.64%) |
| Other/Mixed | 86 (9.74%) | 185 (9.44%) | 136 (12.90%) | 364 (9.91%) |

Note. Ext = Externalizing disorders without comorbidity, Int = Internalizing disorders without comorbidity, Com = Comorbidities between internalizing and externalizing disorders, HC = healthy control.

## Measures

### Structural image acquisition and quality control

T1-weighted structural MRI data were gathered on 3T MRI systems (Siemens Prisma, General Electric MR 750, Philips). On the basis of standardized processing pipelines (26), structural MRI data processing were collected using FreeSurfer version 5.3.070. All scan sessions completed radiological review whereby scans with incidental results were identified. Participants were removed who could not pass visual inspection of T1 images and FreeSurfer quality control (40) (imgincl_t1w_include==1). According to the Desikan-Killiany Atlas, The current analysis used post-processed SA and CT data which were mapped to 34 cortical parcellations per hemisphere (68 brain regions in total) (41).

### Genetic data

We carryed out genotype imputation using high-quality genotyped data made up of 516,598 single nucleotide polymorphism (SNPs) and 11,099 individuals by IMPUTE2 software. The imputation reference set is from the Phase 3 of 1000 Genomes Project. 3 factors are included in Post-imputation quality control: Minor Allele Frequency <0.5%, or out of Hardy-Weinberg equilibrium violation (p>10^−6^), SNPs with imputation info score<0.7, >10% missing rate and individuals with >10% missing rate. We utilize plink v2.0. GWAS to calculate genetic relatedness and only 4,933 European ancestry whose genetic ancestry factor of European>0.9 and 8,498,283 SNPs (kinship coefficient<0.125) are included in subsequent analysis. We performed genetic Principal Component Analysis on unrelated European individuals and derived first ten genetic principal components to correct for population stratification.

### Child Behavior Checklist (CBCL)

The Child Behavior Checklist (CBCL), collected by the child’s caregiver or parents, is generally used to measure emotional and behavioral problems for children. The resulting scores used in ABCD include eight syndrome scale scores (Anxious/Depressed, Withdrawn/Depressed, Somatic Complaints, Social Problems, Thought Problems, Thought Problems, Rule-Breaking Behavior, Aggressive Behavior), three summary scores (Internalizing Problems, Externalizing Problems and Total Problems), six DSM-oriented scale scores (Depressive Problems, Anxiety Problems, Somatic Problems, Attention Deficit/Hyperactivity Problems, Oppositional Defiant Problems and Conduct Problems) and three 2007 Scale Scores (Sluggish Cognitive Tempo, Obsessive-Compulsive Problems and Stress Problems). In the current analyses, we used raw scores of 16 CBCL scales from the baseline (n=11,870).

### Genotyping and imputation

Saliva or whole blood samples of participants were collected at the baseline visit and sent from the collection site to Rutgers University Cell and DNA Repository for storage and DNA isolation. Genotyping was performed using the Smokescreen array (42) containing 733,293 SNPs. Quality control (QC) for genotyped data was performed, resulting in 11,099 individuals and 516,598 SNPs. We derived genetic ancestry using fast Structure (43) and genetic relatedness and principal components (PC) using plink v2.0. Genotype imputation was performed on high-quality variations using the IMPUTE2 software and the imputation reference set was obtained from the Phase 3 of 1000 Genomes Project. After imputation, the individuals with >10% missing rate and SNPs with imputation info score<0.7, >10% missing rate, Minor Allele Frequency (MAF) below 0.5%, or out of Hardy-Weinberg equilibrium violation (p>10^−6^) were excluded. Only European ancestry (genetic ancestry factor of European > 0.9) and genetic unrelated (kinship coefficient<0.125) participants were included in the following genetic analysis. The final European genotyped data included 4,933 unrelated individuals and 8,498,283 SNPs. We performed genetic Principal Component Analysis (PCA) to derive first 10 principal components (PC).

### Case-control analysis and ANOVA

We employed linear mixed models (LMM) with the MATLAB (R2018b) to estimate the difference in CT and SA among each of three transdiagnostic groups (externalizing, internalizing and comorbid) to the healthy children group. Our LMM included random effects for family nested within acquisition site. At the same time, it included fixed-effect covariates for sex, age, race/ethnicity (White, Hispanic, Black, Asian, Other/Mixed), pubertal status, parental marital status, total intracranial volume, parental education and body max index (BMI). All analyses were False Discovery Rate (FDR, q=0.05) corrected for multiple comparisons. We also implemented an Analysis of Variance (ANOVA) to examine the difference in CT and SA among externalizing, internalizing disorders, comorbidity and the healthy preadolescents after regressing out the same covariates using LMM. Tukey Test was also performed among four groups in Post hoc analysis.

### Correlation with symptoms

A general psychopathology factor (p-factor) and three sub-factors, externalized disorder (EXT), internalized disorder (INT) and thought disorder (THO), were modeled using the parent-rated K-SADS-5 (44, 45). Based on a prior observation from the ABCD study (36), employing a hierarchical model of externalizing (ADHD, ODD, CD), internalizing (MDD, GAD, PTSD, PD, SEP, SAD), thought (hallucinations, delusions, OCD, BP) disorder scores, we derived the p-factor using confirmatory factor analysis (R v4.0, *cfa* function of *lavaan* package). This analysis was based on the whole sample (N = 11,878). We also performed association analyses between total CBCL scores and the morphometric variables SA extracted from regions affected in the comorbid group compared to healthy subjects. For the CBCL symptom correlations, all children with symptom scores (n=7570) were included irrespective of the diagnostic classifications, after regressing out the same covariates using LMM.

### Genome-wide association study (GWAS)

In order to check if different regions share common genetic variations, we performed GWAS on 29 regions for SA and 15 brain regions for CT, that had significant SA and CT changes in at least one diagnostic family compared to healthy control subjects.

Genomic risk loci were defined using the FUMA (46) online platform (version 1.3.6a). Independent significant SNPs (IndSigSNPs) were defined as variants with a P value< 5×10^−8^ and independent of other significant SNPs at r^2^<0.6. Lead SNPs were also identified as those independent from each other (r^2^<0.1). LD blocks for IndSigSNPs were then constructed by tagging all SNPs with MAF ≥0.0005 and in LD (r^2^≥0.6) with at least one of the IndSigSNPs. Reference panel population was European of the 1000 Genomes Project Phase 3. IndSigSNPs and all of the tagged SNPs were subsequently searched by FUMA in the NHGRI-EBI GWAS catalog (47) (https://www.ebi.ac.uk/gwas/) to find their previously reported associations (P<0.05) with any traits. All p-values passed FDR correction (FDR q=0.05).

On the one hand, SNPs were mapped to genes by a combination of positional, eQTL and 3D Chromatin Interaction mappings. Specifically, positional mapping was mapping SNPs to locus based on their physical positions. In eQTL mapping, SNPs was mapped to candidate genes according to significance criteria (P<0.05) eQTL associations from several data resources of eQTLs, such as GTEx (48), BRAINEAC (49) and CommonMind Consortium (50). All p-values passed FDR correction (FDR q=0.05). On the other hand, to combine cumulative effects of SNPs assigned to a gene, gene-based analysis was carried out using MAGMA (51) implemented in FUMA. SNPs were mapped to genes with no window around genes (window size=0). Then the gene-based P values were calculated by the GWAS summary statistics of mapped SNPs, indicating the association between the gene and the GWAS phenotype. Genes significantly associated with each of ROI with diagnostic effect on SA and CT were determined by FDR correction (FDR, q=0.05).

### Gene set enrichment analysis

To further identify the biological processes underlying regional SA and CT, we performed gene set enrichment analyses on regional SA and CT based on KEGG, GO and GWAS catalog gene sets. All genes were set as background genes. FDR (FDR, q=0.05) correction for all analyses was applied through FUMA. Other parameters in these analyses were set as default.

### Cell type specificity analysis

To test whether genetic risk variants for regional SA and CT converge on a specific cell type, we performed cell type specificity analysis (52) using 7 single-cell RNA sequencing datasets from human brain tissue (Supplemental Table S1) and pre-computed MAGMA results, which builds the relationships between cell type-specific gene expression and trait–gene associations. We used False Discovery Rate (FDR, q=0.05) correction for multiple testing in per dataset to identify significantly associated cell types.

## Results

### Surface area: more brain regions affected in comorbidity than single diagnostic families

Children with comorbidity had pronounced SA reductions across the brain compared to the controls, while the single diagnostic groups only had a few regions with significant changes compared to the controls (Figure 2). In particular, 29 out of 68 cortical regions demonstrated significant decrease in SA in comorbid children, including the left precuneus, right middle temporal gyrus, left supramarginal, and prefrontal areas (left pars orbitalis, right pars orbitalis) and sensory motor regions (right postcentral gyrus, left precentral gyrus), all p-values passed FDR correction (FDR q=0.05), see Supplement Table S2. When children with externalizing disorders were compared with healthy children, only 2 temporal regions (right inferior temporal gyrus and left superior temporal gyrus) demonstrated a significant SA reduction, while no SA reduction was notable in the internalizing disorder group (Figure 2 and 3). The two temporal regions with reduced SA in externalizing disorder group also showed SA reduction in the comorbid group.

**Figure 2.**
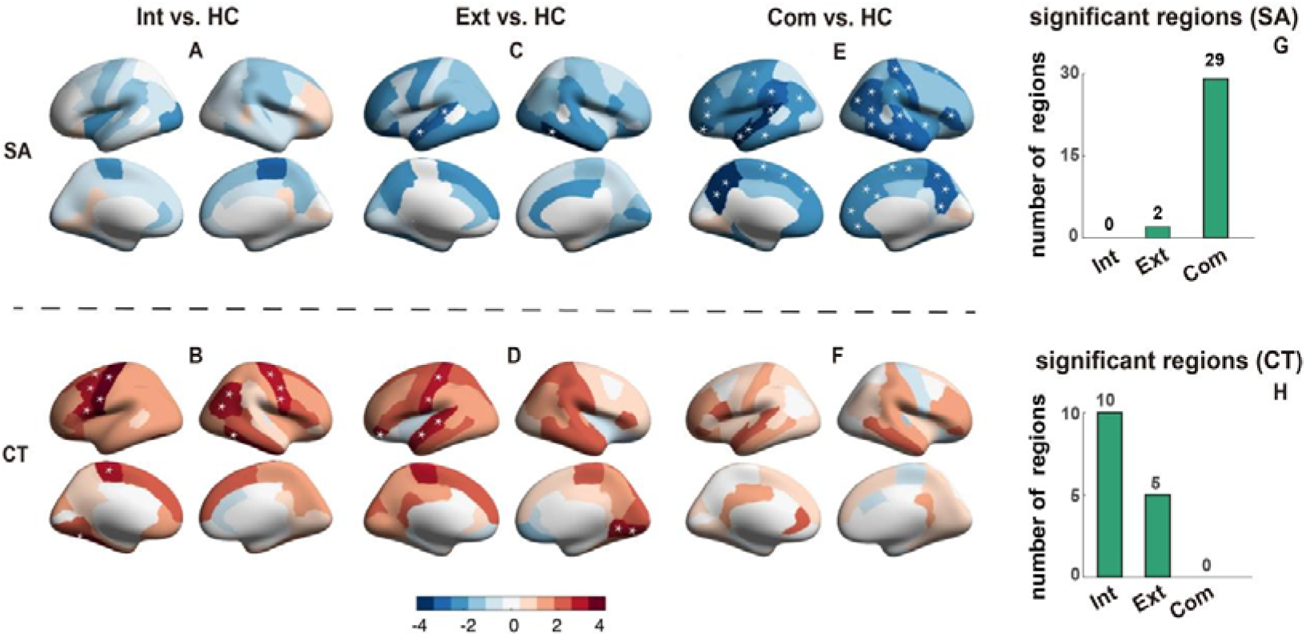
Brain regions with significant morphological alterations compared to healthy control group in externalizing disorders group, internalizing disorders group and the comorbidity group. Brain regions with significant morphological difference compared to healthy control group in externalizing disorders group **(A, B)**, Internalizing disorders group **(C, D)** and Comorbidity **(E, F)** in terms of cortical surface area **(A, C, E)** and cortical thickness **(B, D, F)**. The color bars in **(A-F)** represent the t value of the regression coefficient of group variable from Linear Mixed Model (LMM). The regions with * represents P<0.05, FDR corrected (FDR q=0.05). The number of brain regions with significant changes for each of three transdiagnostic groups (externalizing, internalizing and comorbidity group) are shown for cortical surface area **(G)** and cortical thickness **(H)**. Abbreviations: Int, Internalizing disorders; Ext, Externalizing disorders; Com, Comorbid between internalizing and externalizing disorders; HC, healthy control.

**Figure 3.**
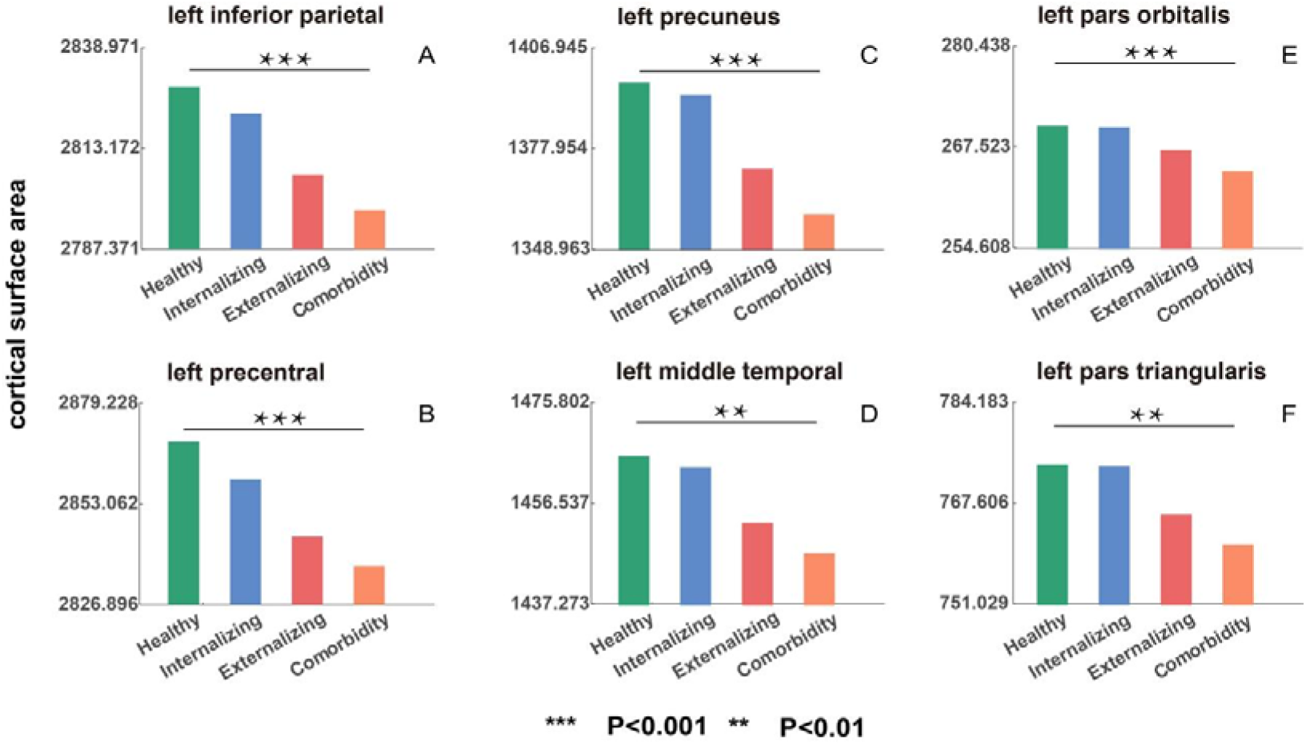
The comparison of mean cortical surface area (SA) in controls, internalizing, externalizing and comorbidity group. Mean SA of controls, internalizing, externalizing and comorbidity group for regions with significant SA changes in comorbidity group (compared to controls). Only the top six regions with significant SA alterations in comorbidity group were shown. Statistics of more brain regions can be found in Supplement Table S2. The y axis represents the mean SA. All p-values passed FDR correction (FDR q=0.05).

ANOVA analysis contrasting the 4 groups (internalizing, externalizing, comorbidity disorder and control group) for SA revealed significant brain regions mainly including left supramarginal gyrus, right pars orbitalis, right postcentral gyrus and left precentral gyrus, in accordance with the above case-control results (Supplement Table S3). A post hoc head-to-head contrast revealed SA reduction affecting left precuneus and right pars triangularis in the comorbid group compared to the internalizing disorders and the control group, though no notable differences in SA was observed in the comorbid group in direct comparison with the externalizing disorder group. Taken together, these observations indicate that the SA changes in comorbidity not only includes those that are seen in externalizing disorder, but also exceed them in magnitude; further, SA changes in the comorbid group are more extensive than the minimal, insignificant deviations seen in internalizing disorders. This is also reflected in figure 3 in which comorbid group is more “similar” to the externalizing disorder group than the internalizing disorder group.

### Cortical thickness: more regions affected in single diagnostic families than comorbidity group

Children with either internalizing or externalizing disorders had significant alterations in CT when compared to healthy children (externalizing disorders: 5 regions, internalizing disorders: 10 regions, see Figure 3). The comorbid group had no significant differences in CT in any of the 68 brain regions compared with the healthy children. For externalizing disorders family, auditory (left transverse temporal gyrus and left superior temporal gyrus), sensory-motor (left postcentral gyrus), visual (right lingual) and prefrontal cortex (left pars orbitalis) showed significant CT increases. For internalizing disorders, sensory-motor (left precentral gyrus, right precentral gyrus and left paracentral lobule), temporal (right inferior temporal gyrus and right banks of superior temporal sulcus) and frontal-parietal (left pars opercularis, left caudal middle frontal gyrus, right inferior parietal gyrus and left superior frontal gyrus) cortices showed significant increases (Supplement Table S4).

ANOVA contrasting the 4 groups (internalizing, externalizing, comorbidity disorders and control group) revealed significant changes in line with the above analysis, with changes affecting bilateral precentral gyrus, left pars opercularis, temporal cortex (right inferior temporal gyrus and right banks of superior temporal sulcus) and right inferior parietal gyrus, see Supplement Table S5. A post hoc head-to-head contrast revealed higher CT in bilateral paracentral lobules in children with externalizing disorders, compared to both the healthy and the comorbid group. Children with internalizing disorders had higher CT in the left paracentral lobule and bilateral precentral gyrus compared to the other 2 groups. Taken together, these observations indicate that the CT changes in internalizing and externalizing disorders are extensively and variously distributed, but in the presence of comorbidity these changes do not co-occur; Instead they diminish in magnitude, leading to a pattern that is indistinguishable from healthy controls. Finally, ANOVA analysis show that p values are less significant when comparing comorbid group to externalizing disorder group than internalizing disorder group, suggesting that comorbid group is more “similar” to the externalizing disorder group (Supplement Table S5).

### Surface area but not cortical thickness satisfies additivity of effects

We found that alterations of surface area for externalizing and comorbid (internalizing and externalizing) group reflect an additive effect. In other words, the SA changes in the comorbid state appear to be partially an aggregate of the individual changes that occur in internalizing and externalizing disorders. In contrast, CT in the comorbid group does not satisfy the expectations under an additive model, as internalizing/externalizing group each had significant CT increase (compared to controls) but the comorbid group did not have more pronounced CT increase, as one would expect under the additive model; instead, comorbidity was related to lack of changes in CT compared to healthy controls.

### Psychiatric symptom measures correlated with surface area but not cortical thickness

We found that p-factor, which reflects an overarching susceptibility to any mental disorder (44, 53, 54) was significantly higher in children with a notable reduction in the SA (irrespective of diagnostic status) in the cortical regions that were prominently affected in the comorbid group (bilateral precuneus, superior and inferior temporal gyrus, and the left superior frontal gyrus; all P<0.05, FDR corrected (FDR q=0.05), see Figure 5A). However, p-factor was not correlated with cortical thickness (Figure 5B).

**Figure 4.**
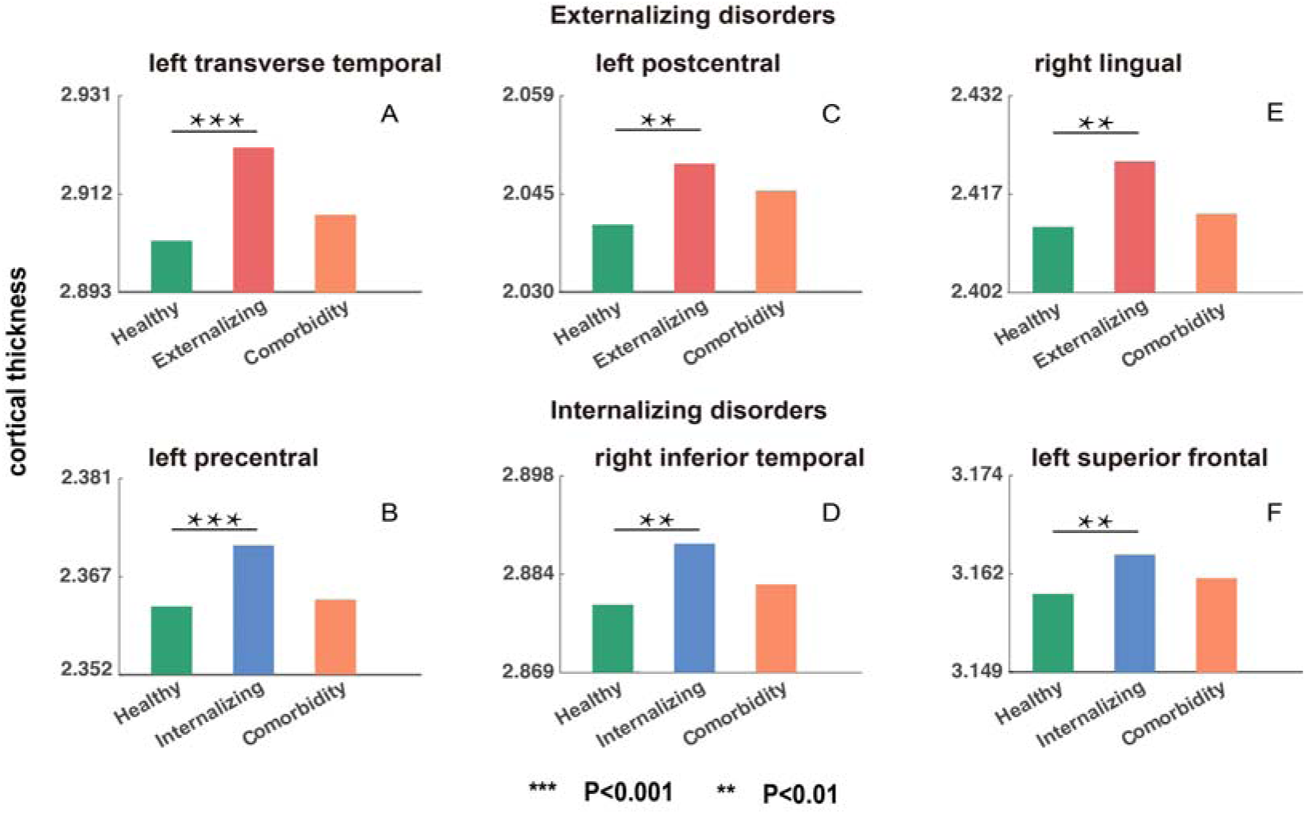
The comparison of mean cortical thickness (CT) in controls, internalizing, externalizing and comorbidity group. Mean CT of controls, externalizing, and comorbidity group for regions with significant CT changes in externalizing group (compared to controls) **(A, C and E)**. Mean CT of controls, internalizing, and comorbidity group for regions with significant CT changes in internalizing group (compared to controls) **(B, D and F)**. Only the most significant 3 regions were shown for each disorder, and the rest brain regions were shown in Supplement Table S4. The y axis represents the mean CT. All p-values passed FDR correction (FDR q=0.05).

**Figure 5.**
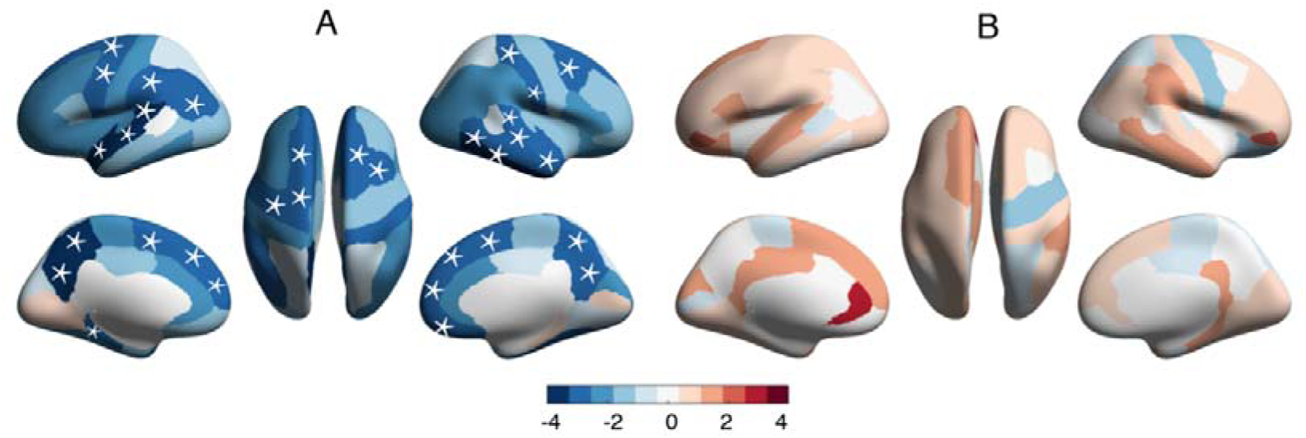
Correlation between p-factor reflecting an overarching susceptibility to any mental disorder and SA and CT. Correlation between p-factor and SA in comorbidity-specific regions **(A)**, and correlation between p-factor and CT in externalizing-specific regions and internalizing-specific regions **(B)**. The color bar represents the t value of the regression coefficient of group variable from Linear Mixed Model (LMM). The asterisks (*) indicates P<0.05, FDR correction (FDR q=0.05).

Children with lower SA in the cortical regions affected by comorbidity also had higher CBCL Externalizing Problems scores (including Rule-Breaking Behavior scores, Aggressive Behavior scores, Oppositional Defiant Problems scores and Conduct Problems scores) and Internalizing Problems scores (including Withdrawn/Depressed scores and Depressive Problems scores) with P<0.05, FDR corrected (FDR q=0.05), see Figure 6. In particular, the SA of prefrontal and temporal regions relate to both Externalizing and Internalizing Problem scores.

**Figure 6.**
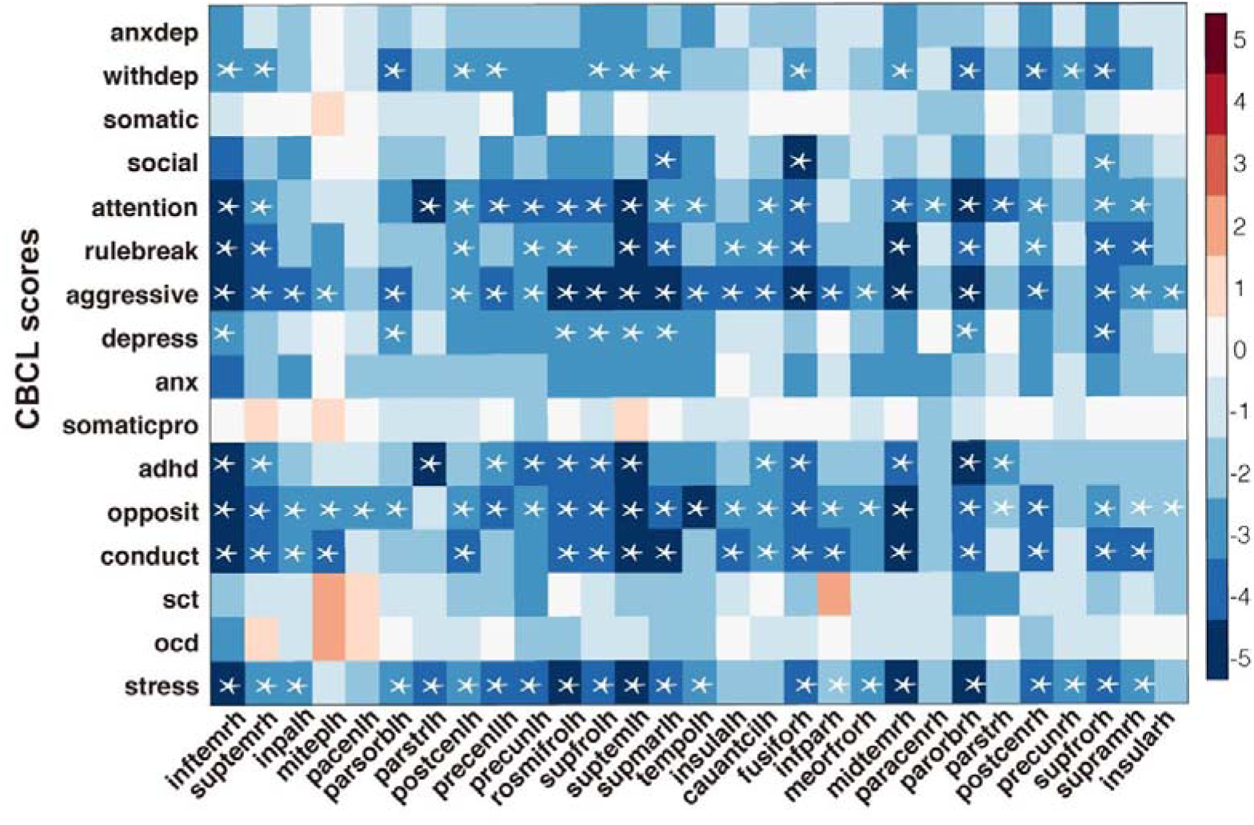
Associations between CBCL score and surface area of regions with significant alterations in patient groups. The color bar represents the t value of the regression coefficient from LMM. The asterisks (*) indicates P<0.05, FDR correction (FDR q=0.05). Abbreviations: AnxDep, Anxious/Depressed; WithDep, Withdrawn/Depressed; Somatic, Somatic Complaints; Social, Social Problems; Thought, Thought Problems; Attention, Attention Problems; Rulebreak, Rule-Breaking Behavior; Aggressive, Aggressive Behavior; Anxdisord, Anxiety disorders; Somaticpr, Somatic Problems; Opposite, Oppositional Defiant Problems; Conduct, Conduct Problems; OCD, Obsessive-Compulsive Problems; Stress, Stress Problems; ADHD, Attention Deficit/Hyperactivity Problems; picvocab, Picture Vocabulary; inftemrh, right inferior temporal gyrus; suptemrh, right superior temporal gyrus; inpalh, left inferior parietal gyrus; miteplh, left middle temporal gyrus; pacenlh, left paracentral lobule; parsorblh, left pars orbitalis; parstrlh, left pars triangularis; postcenlh, left postcentral gyrus; precenllh, left precentral gyrus; precunlh, left precuneus; rosmifrolh, left rostral middle frontal gyrus; supfrolh, left superior frontal gyrus; suptemlh, left superior temporal gyrus; supmarlh, left supramarginal; tempolh, left temporal pole; insulalh, left insula; cauantcirh, right caudal anterior cingulate; fusiforh, right fusiform; infparh, right inferior parietal gyrus; meorfrorh, right medial orbito frontal gyrus; midtemrh, right middle temporal gyrus; paracenrh, right paracentral lobule; parorbrh, right pars orbitalis; parstrh, right pars triangularis; postcenrh, right postcentral gyrus; precunrh, right precuneus; supfrorh, right superior frontal gyrus; supramrh, right supramarginal; insularh, right insula.

### Distinct biological process and cell types associated with SA and CT alterations

In order to understand the genetic underpinnings of the difference between SA and CT alterations across single diagnostic families (internalizing or externalizing disorders) and the comorbid group, we performed GWAS, gene set enrichment analysis and cell type specificity analysis. We first performed GWAS for the 29 ROIs with significant SA changes and 15 ROIs with significant CT changes in the patient groups, using 4,716 European-ancestry unrelated individuals. 19 and 8 regions showed significant associations for SA and CT, respectively (p< 5 × 10^−8^, Supplement Table S6-32). All p-values passed FDR correction (FDR q=0.05). Next, SNPs were mapped to genes with a combination of positional, eQTL and 3D Chromatin Interaction mappings by FUMA. MAGMA gene-based association also identified several significantly associated genes in FUMA. Then, we performed enrichment analyses using all the above genes.

The major biological pathways that relate to SA differ from those related to CT. Although both modalities are associated with neurogenesis, SA-related genes are enriched primarily in protein phosphorylation, while CT-related genes are primarily enriched in central nervous system development, positive regulation of developmental process and cell differentiation (Supplement Table S33-34). Furthermore, in the gene set enrichment analysis, distributed CT alterations were associated with common genes relating to immune-related biological processes (Supplement Table S35). For example, TNIP3 and PRDM5 genes were linked to CT of bilateral precentral gyrus; MCM2, PODXL2 and MGLL to left postcentral and left precentral regions; all of these genes being linked to immune-related biological processes. MCM2 and PODXL2 are associated with oligodendrocytes (55–57), and MCM2 has a regulatory role in a range of oligodendrogliomas (57), and the PODXL2 gene is over expressed in oligodendrocytes in zebrafish (56). TNIP3 is involved in the regulation of interferon IFN-β1 that is pro-inflammatory (58, 59).

Finally, cell-type specificity analysis for the genes associated with SA or CT alterations (Supplement Table S36-37) uncovered further differences. SA-related genetic pathways specifically relate to the excitatory neurons while CT-related cell types include microglia and excitatory pyramidal neurons. For SA, genes related to regions most affected in comorbid group (left rostral middle frontal gyrus) had significant associations with Exc_L2.3, Exc_L2.4, Exc_L3.4, Exc_L3.5, Exc_L4.5, Exc_L5.6 and Exc_L6 (excitatory neurons). For CT, three internalizing-specific regions (right precentral, left pars triangularis and right ITG) and one externalizing-specific region (left postcentral) all showed significant associations with microglia. Moreover, an externalizing-specific region (left SFG) showed significant associations with exCA1 and exCA3 (excitatory pyramidal neurons in the hippocampal Cornu Ammonis region).

## Discussion

In a large preadolescent sample of 1,878 children, we studied the structural basis of comorbid expression of internalizing and externalizing disorders and reported 2 major findings. First, children with comorbidity show more pronounced deviation from healthy children in cortical SA across fronto-temporal cortex. This reduction is much more pronounced than what is seen in children with externalizing disorder who in turn show more reduction than those with internalizing disorders. The magnitude of SA reduction also tracks the severity of the ‘p’ factor. Second, children with comorbidity show near-normal CT. This is in contrast with the aberrant increase in CT seen in children with internalizing disorders, who show more pronounced increase compared to those with externalizing disorders. Interestingly, increase in CT does not relate to the ‘p’ factor reflecting comorbid psychopathology. Therefore an interesting gradient was observed in both CT and SA: the degree of changes in externalizing disorders was closer in magnitude to comorbidity, than internalizing disorders for both SA and CT. This is also supported by the findings that a higher percentage of externalizing children converted to comorbidities after two years compared to internalizing children (Supplement Table S38). Taken together, our findings indicate a specific role for the maturation of SA in the development of comorbid disorders, while the mechanistic pathways underlying individual diagnostic families likely operate via distinct aberrations in CT.

Cortical SA changes correlated significantly with the p-factor which reflects an overarching susceptibility to several mental disorders (44, 53, 54), see Figure 5A. This is consistent with previous studies on the ABCD cohort (22, 37). Further, across the frontotemporal regions affected in the comorbidity group, SA significantly related to both Internalizing and Externalizing Problem scores (Figure 6). This again reinforced the suggestion that SA, rather than CT, underlies the emergence of a comorbid disorder pattern, and this may relate to the continuous nature of the relationship between SA and psychopathology in this age group.

The distinct patterns of structural alterations in SA and CT are in line with the fact that these two morphological measures are genetically independent (27, 28, 30). Using cell-type specificity analysis of genes associated with regional SA and CT alterations (Supplement Table S38-39) we parsed this further. For left rostral middle frontal gyrus, a region with reduced SA in the comorbid group, we noted significant associations especially with C1ql3-expressing excitatory neurons. C1ql3-deficient mice have been shown to exhibit fewer excitatory synapses and diverse behavioral abnormalities such as fear memory impairments (60). For CT, three internalizing-specific regions (right precentral, left pars triangularis and right ITG) and one externalizing-specific region (left postcentral) all showed significant associations with microglia, the main immune cell in the brain (22, 37, 61, 62). Microglia has been shown to play a role in synaptic plasticity, neurogenesis and memory (63, 64) and various psychiatric disorders in adolescence (65, 66). Taken together, a generalized vulnerability affecting excitatory synapses and thus cortical surface area may underlie preadolescent mental disorders. In children with higher doses of this vulnerability, SA reduction is more pronounced and comorbid diagnostic states are expressed. An independent, immune mediated pathway also operates in children with psychopathology, though not directly contributing to comorbidity.

We note that the regions with significant CT alterations have common genetic associations relating to immune-related biological processes (Supplement Table S37), supporting a possible role for the neuroimmune system in CT alterations underlying psychopathology. Given the broad stress-resilience role played by the immune system, these findings indicate a mechanism by which diathesis-stress interplay can operate at the cortical structure level (67), possibly mediated by the microglia (66, 68–71). Stress has been shown to be associated with increased pro-inflammatory cytokines and excessive inflammation for children (72, 73), leading to microglia activation (69) and reduction in neuronal/synaptic density (66). Another possibility, highlighted by the link to oligodendrocyte-related genes, is that increased CT may relate to delayed or insufficient myelination resulting in a poorly defined gray–white boundary (74) and therefore thicker gray matter (75) in children with internalizing or externalizing disorders. Quantitative myelin mapping studies are required to confirm this notion in the future.

Therefore, we postulate that that there are two immune-modulated processes that caused the observation that significant CT alterations occurred in single disorder but not in comorbidity group. These two processes are related to oligodendrocytes and microglia, respectively, which are the two main constituting parts in the immune system of brain, shown to be able to affect CT in opposite ways (70, 75):

1. Oliodendrocytes: oliodendrocytes are responsible for myelination (76). For children with depression and ADHD, oliodendrocytes have been shown be impaired or reduced in number (77–80) (associated with pro-inflammatory cytokines (81)), leading to weak myelination. Recently it has been shown that voxels with less myelin near the gray–white matter boundary will appear darker in children than adults, which shifts the apparent gray–white boundary deeper into the white matter (74) and thus leads to thicker gray matter (75). Therefore, children with internalizing/externalizing disorder may have weak myelination thus increased CT than controls.
2. Microglia: Microglia plays a physiological role in psychiatric disorders like depression and ADHD during development (82, 83), in which neuronal–microglial interactions are disrupted, resulting in prolonged/increased activation of microglia (69, 84). Stress has been shown to be associated with increased pro-inflammatory cytokines and excessive inflammation for children (72, 73), leading to microglia activation (69) and microglia’s engulfment of a large number of neurons (66). As comorbidity group’s stress level is more severe than both internalizing/externalizing group and control group, it results in excessive microglial activation and possibly microglial engulfment of neurons (66, 68–71) thus decreased CT.

Therefore, oliodendrocytes dysfunctions and microglia overactivation leads to increased and decreased CT, respectively, all related to level of pro-inflammatory markers. As we have shown that stress level increases from controls to single diagnostic families to comorbidity group, and that stress level also correlated positively with the pro-inflammatory markers (interleukin (IL)-1β and IL-6) in psychiatric disorder (61, 62), pro-inflammatory markers thus are also expected to increase from controls to single diagnostic families to comorbidity group. Therefore for single diagnostic families with increased pro-inflammatory markers, oligodendrocyte impairments play a major role and leads to increased cortical thickness. For the comorbidity group in which stress and pro-inflammatory factor are further increased, microglia overactivation is present, leading to neuron engulfment and likely thinner cortical thickness, counteracting the increased thickness seen in single diagnostic families. Thus comorbidity group does not show significant CT changes.

In a broad sense, if we assume that a continuously distributed neurobiological mechanism underlies both internalizing and externalizing disorders, then that risk is likely reflected in the development of SA, with the most affected individuals displaying the most severe SA reduction, and internalizing disorder being the least affected. Similarly, if we construe CT changes to reflect resilience in response to this underlying continuous dimension of risk, then higher CT may confer higher degree of resilience, with the most resilient one displaying internalizing phenotype, and the least resilient developing notable comorbidity. The immune-related links to CT, rather than SA, support this risk-adaptation model.

## Limitation

Our study has a number of strengths as well as limitations. We examined comorbidity in one of the largest developmental neuroimaging cohorts studied to date; we used multilevel analysis linking genetic variants, cell-types and distinct morphometric variables. Nevertheless, we lacked direct measures of myelination or microglial activity to infer the mechanistic processes in CT alterations in more detail. We also lacked sufficient data to resolve the temporal relationship between brain-based metrics and the behaviors of interest. More direct evidence and verification are needed in future analysis.

## Conclusions

In this large cross-sectional observation, presence of comorbid psychopathology related strongly to reduced SA while higher CT was distinctly limited to children without comorbidity. The proximity of morphometric patterns in comorbid cases to externalizing, rather than internalizing disorder, supports the proposal that externalizing problems increase the secondary risk for internalizing problems at least in some patients, thus resulting in comorbidity. A specific developmental pathway involving excitatory neurons and synapses that operate via areal expansion emerges as a strong candidate risk mechanism for comorbidity.

## Data Availability

LP reports personal fees from Janssen Canada, Otsuka Canada, SPMM Course Limited, UK, Canadian Psychiatric Association; book royalties from Oxford University Press; investigator-initiated educational grants from Janssen Canada, Sunovion and Otsuka Canada outside the submitted work. All other authors report no biomedical financial interests or potential conflicts of interest. None of the above-listed companies or funding agencies have had any influence on the content of this article.

## Acknowledgments

JZ was supported by Shanghai Municipal Science and Technology Major Project (No.2018SHZDZX01) and ZJLab and NSFC 61973086. JF was supported by the 111 Project (No. B18015), the key project of Shanghai Science and Technology (No. 16JC1420402), National Key R&D Program of China (No. 2018YFC1312900), National Natural Science Foundation of China (NSFC 91630314). LP acknowledges salary support from the Tanna Schulich Chair of Neuroscience and Mental Health. Kai Zhang is Sponsored by Shanghai Pujiang Program.

