## Supplemental materials for "Specificity of cortical area and thickness as biomarkers for comorbid internalizing and externalizing mental disorders in pre-adolescence"

### P factor

A general psychopathology factor and three sub-factors, externalized disorder, internalized disorder and thought disorder, were modeled using the parent-rated K-SADS-5. Based on a previous literature that also used data from ABCD study (1), a hierarchy model including externalizing (ADHD, ODD, CD), internalizing (MDD, GAD, PTSD, PD, SEP, SAD), and thought (hallucinations, delusions, OCD, BP) disorder pathology, as well as a p factor using confirmatory factor analysis (R v4.0, cfa function of lavaan package). The analysis was based on the whole sample (N = 11,878) and the final factor scores were used in association analyses.

### Conversion rate between single diagnostic families and comorbidity group

We used 2-year follow-up ABCD data (474 externalizing disorders, 982 internalizing disorders, 541 comorbidities between internalizing and externalizing disorders) to evaluate the conversion rate between single diagnostic families (internalizing disorders and externalizing disorders) and comorbidity group. The number of each diagnostic family after two years divided by the number of each diagnostic family at baseline is the conversion rate.
